# Episia: An Open-Source Python Library for Epidemiological Surveillance, Modeling, and Biostatistics in Resource-Limited Settings

**DOI:** 10.64898/2026.04.17.26350337

**Authors:** Fildouinde Ariel Shadrac Ouedraogo

**Affiliations:** Université Joseph Ki-Zerbo, Faculté de Médecine, Ouagadougou, Burkina Faso; Xcept-Health, Ouagadougou, Burkina Faso

## Abstract

Despite the evolution of epidemiological analysis and modeling tools, difficulties still remain, especially in developing countries, regarding the availability and use of these tools. Often expensive, requiring high technical expertise, demanding constant connectivity of several or sometimes even significant resources, these tools, although efficient, present a major gap with the operational realities of health districts. It is in this context that we introduce Episia, an open-source Python library designed and conceived to provide a framework to facilitate epidemiological analysis and modeling. It integrates a suite of compartmental epidemic models (SIR, SEIR, SEIRD) with a sensitivity analysis using the Monte Carlo method, a complete biostatistics suite validated against the OpenEpi reference standard, as well as a native DHIS2 client for automated data ingestion. Developed in Burkina Faso, it is optimized and aims not only to address these health challenges encountered in Africa but also remains a versatile tool for global health informatics.

## 1 Introduction

Field healthcare professionals in resource-limited settings encounter daily a paradox in which powerful analytical tools exist but remain largely inaccessible. Either costly, technically complex, sometimes dependent on stable internet connectivity, or demanding in terms of hardware resources, these sophisticated software programs are nevertheless deeply out of step with the operational realities of health districts. This disconnect is evident at several levels. Field epidemiologists rely on fragmented workflows involving manual data export from health information systems, cleaning in third-party tools, and then importing into analysis software—a tedious, time-consuming, and error-prone process. Furthermore, the lack of systematic validation of results against recognized references undermines confidence in the analyses produced.

To address these concrete challenges observed and experienced in the day-to-day practice of public health in Burkina Faso we present Episia, an open-source Python library designed for field epidemiology in resource-limited settings. Episia brings together within a unified framework that can be used offline: compartmental epidemic models (SIR, SEIR, SEIRD) with uncertainty analysis through Monte Carlo simulation, a biostatistical suite numerically validated against OpenEpi, and a native client for direct integration with DHIS2, the main health information system used in Africa.

## 2 Related Work and State of the Field

The ecosystem of epidemiological software is varied, but few offer the integration necessary for public health in the field in low-resource settings.

### 2.1 The R Ecosystem

The R programming language has long established itself as the reference in epidemiological research. Although libraries such as epiR [3] offer biostatistical functions and EpiEstim [4] and EpiModel [5] respectively provide specialized functionalities for the estimation of the time-varying reproduction number (*R*_*t*_) and network-based modeling, none natively support DHIS2. This logically makes the deployment of R-based workflows in the field complex.

### 2.2 Python Libraries

While in the Python ecosystem, libraries such as Statsmodels [6] provide general regression techniques, specialized tools like EpiPy or Epilearn [7] focus on mechanistic modeling of epidemics. Despite their power and performance, the latter seem more often intended for academic researchers who possess strong programming skills and neither integrate specific biostatistical measures (for example, stratified Mantel-Haenszel odds ratios, diagnostic test evaluations) nor the automatic report generation features required by field epidemiologists.

### 2.3 The Gap: DHIS2 and Offline Validation

The common feature of existing software lies in the lack of native integration with DHIS2. Analysts must manually export CSV files from DHIS2, clean them in external software, and then import them into tools of their choice, which is tedious, time-consuming, and increases the margin of error. Furthermore, few modern libraries offer systematic numerical validation against established “references” such as OpenEpi [8], which is widely used in epidemiology training. Episia stands out by offering:

- A native DHIS2 client: direct integration via API for automated data retrieval.
- Validated biostatistics: 100% numerical agreement with OpenEpi on key indicators.
- A unified workflow: modeling, statistics, and reporting in a single library.
- An “offline-first” design: no network dependency during execution, which is crucial for remote field missions.

## 3 Methods and Software Architecture

Episia is designed as a Python module (version 3.9+) relying on Python scientific libraries (NumPy, SciPy, Pandas). Its architecture is subdivided into several functional subsystems, coordinated through a high-level API namespace (episia.epi), and represented in the following table.

**Table 1:**
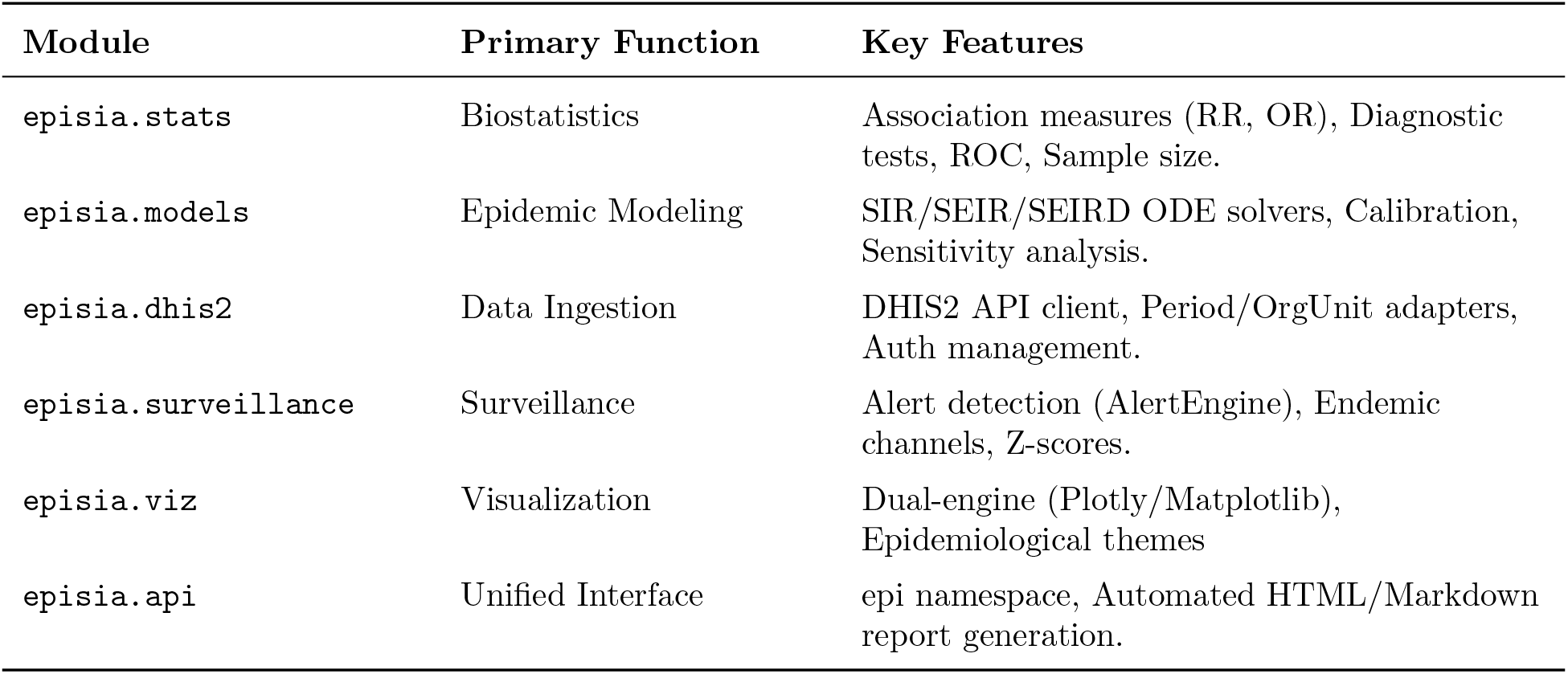
Episia’s Software Architecture, Main Modules.

**Figure 1:**
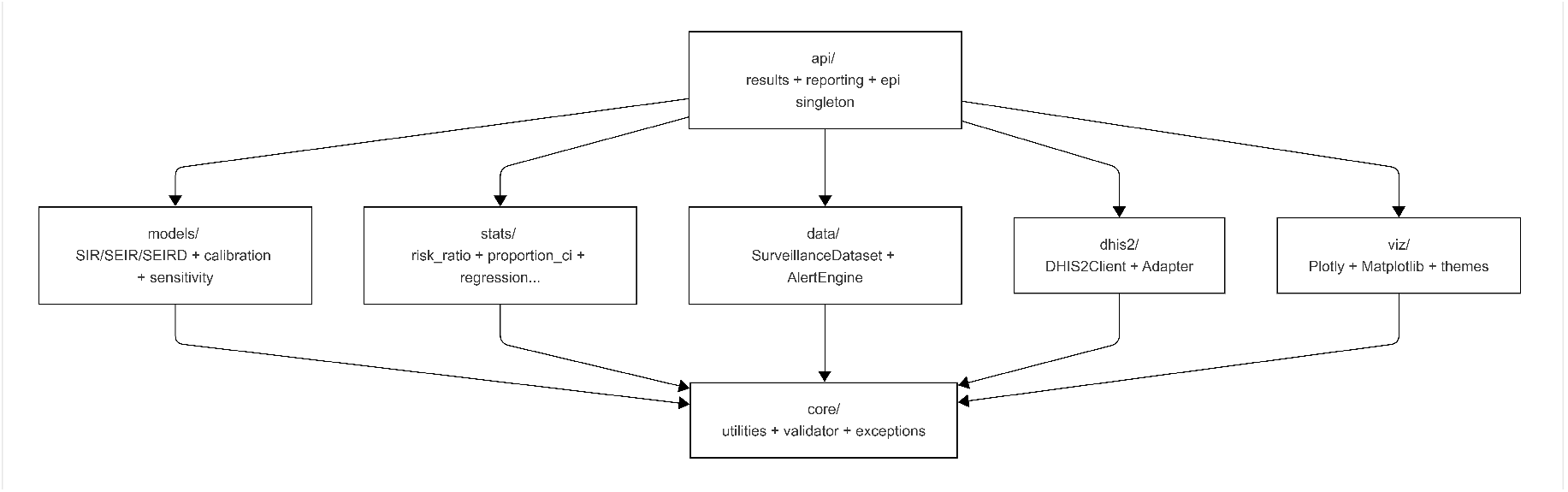
Episia’s Software Architecture Overview.

### 3.1 Biostatistical Engine and Validation

The episia.stats module provides the main statistical methods used in field epidemiology. Beyond point estimates, it offers several confidence interval (CI) methods for proportions (Wilson, Clopper-Pearson, Jeffreys) and means. The major innovation lies in its systematic validation against OpenEpi. We have developed a suite of 1,390 automated tests, including 45 specific “concordance cases” that compare Episia results directly to OpenEpi reference values. Numerical concordance is maintained to at least three decimal places for all validated functions.

### 3.2 Mechanistic Modeling and Uncertainty

The modeling engine solves systems of ordinary differential equations (ODEs) for the SIR, SEIR, and SEIRD compartments. In order to go beyond simple “best-fit” curves, Episia incorporates:

- A calibration system that allows automated adjustment of parameters to observed surveillance data using the L-BFGS-B algorithm
- A sensitivity analysis through Monte Carlo simulations allowing parameters to follow uniform, normal, or triangular distributions, producing “uncertainty envelopes” for epidemic projections
- A scenario comparison through tools that allow the impact of different intervention strategies (for example, social distancing, vaccination) on the epidemic peak and the final epidemic magnitude to be compared.

### 3.3 Surveillance and DHIS2 Integration

The episia.dhis2 module allows analysts to connect directly to HMIS instances. It manages the complexity of the DHIS2 Analytics API, including the conversion of ISO weeks and organizational unit hierarchies. Moreover, the AlertEngine integrated into episia.surveillance implements the WHO standard algorithms for epidemic detection, such as the threshold system of the ‘meningitis belt’ (for example, exceeding 10 or 15 cases per 100,000 inhabitants per week) [2].

**Figure 2:**
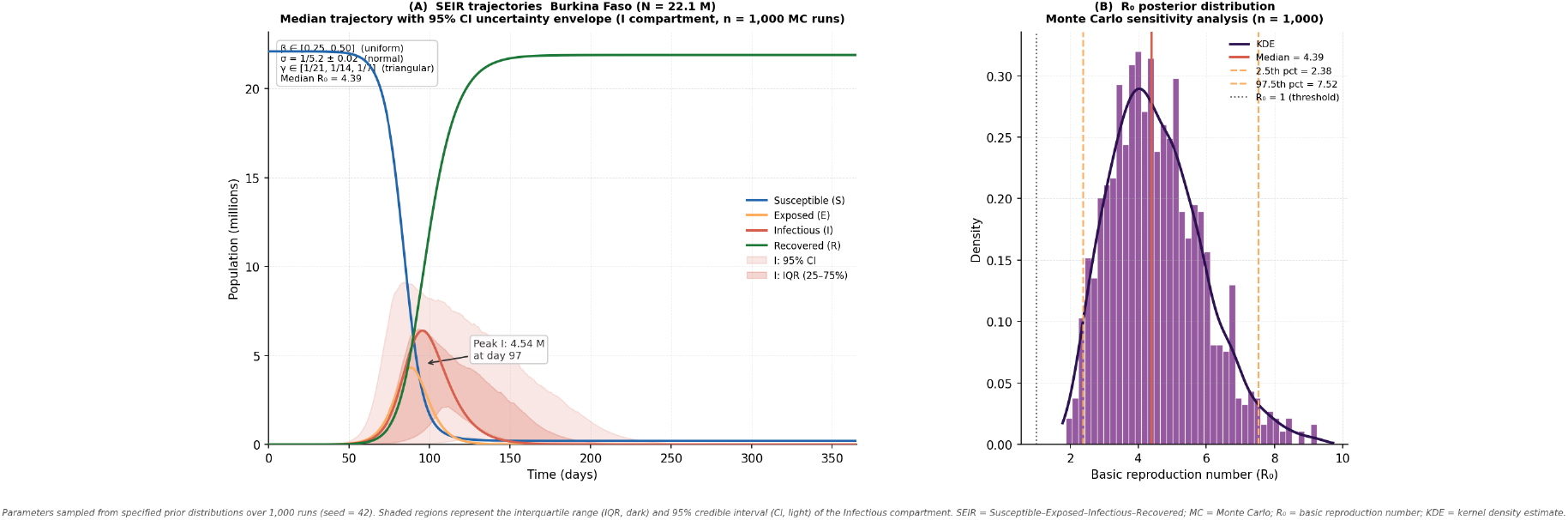
SEIR epidemic trajectories and Monte Carlo uncertainty analysis (Burkina Faso)

**Figure 3:**
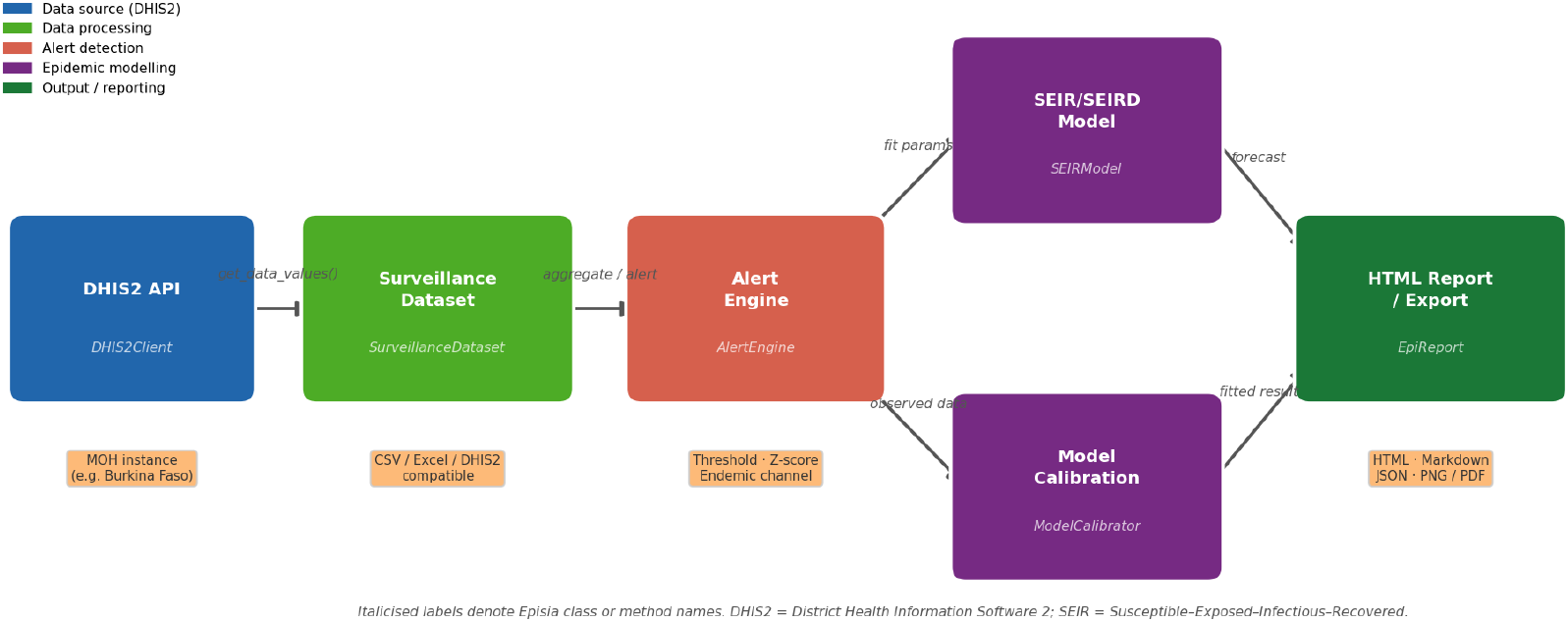
DHIS2 data ingestion and analysis workflow in Episia.

## 4 Validation and testing

We selected OpenEpi [8] as the main reference standard due to its widespread use in epidemiology training and its well-documented, peer-reviewed algorithms. Every major statistical function of episia.stats without exception has been systematically tested against the 14 calculation modules of OpenEpi.

**Table 2:**
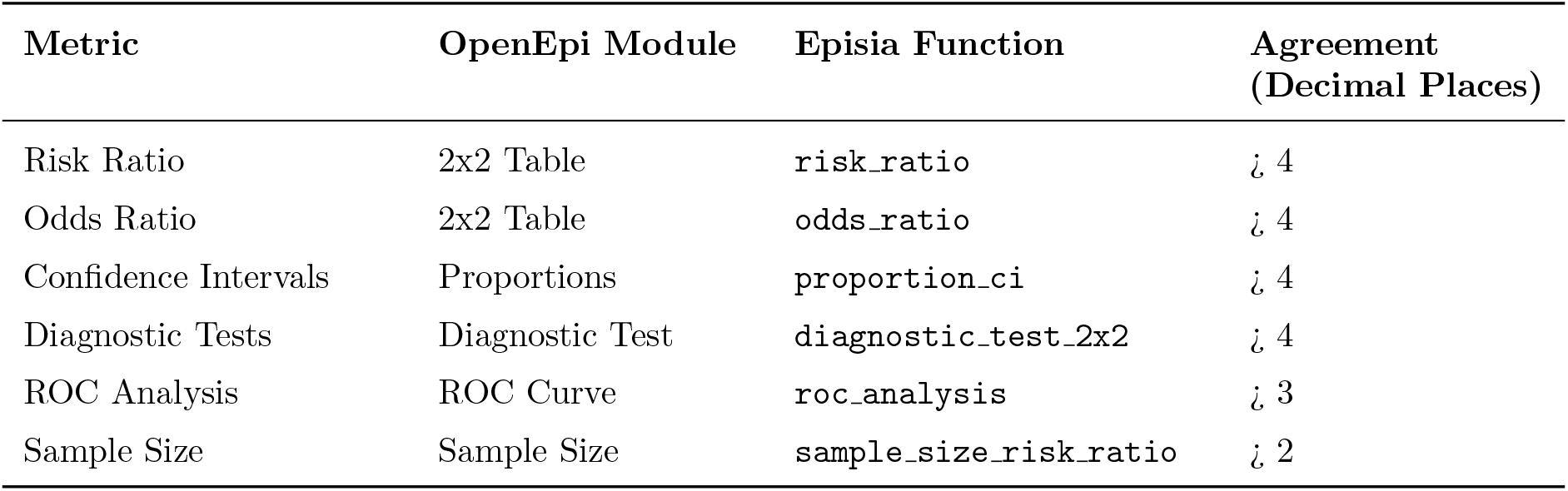
Numerical agreement between Episia and OpenEpi on key biostatistical indicators.

A [validation notebook](https://github.com/Xcept-Health/episia/blob/main/exemples/episia_vs_openepi.ipynb) was set up in order to reproduce the OpenEpi calculations step by step. All 45 independent tests demonstrated numerical agreement, thereby showing that professionals can, just like with OpenEpi results, rely on those of Episia.

**Figure 4:**
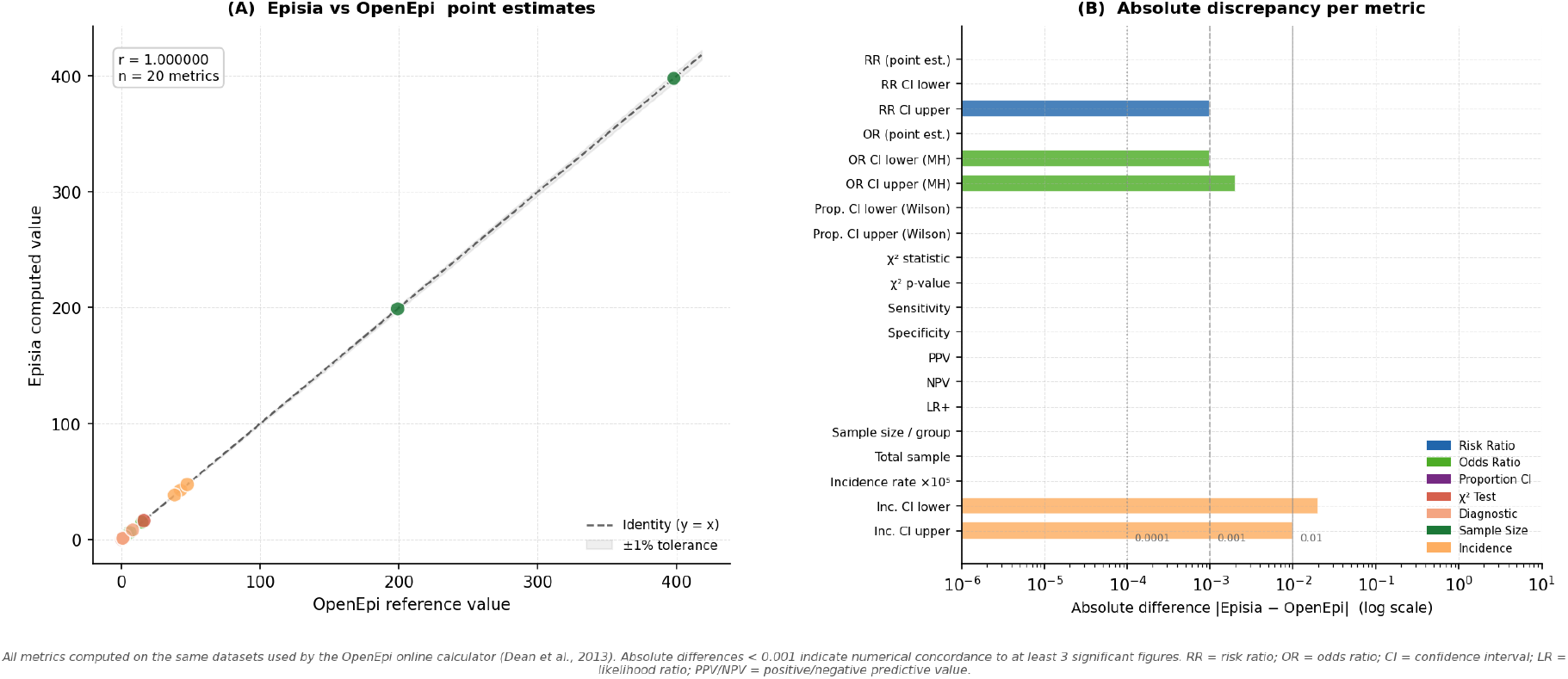
Comparison of Episia outputs with OpenEpi reference calculations.

## Data Availability

All data produced in the present work are contained in the manuscript. The source code and validation notebooks are available at https://github.com/Xcept-Health/episia (DOI: 10.5281/zenodo.19429374).

## 5 Code availability

The source code of Episia is freely available on GitHub at https://github.com/Xcept-Health/episia and archived on Zenodo (DOI: 10.5281/zenodo.19429374). The library is also distributed via PyPI (install with `pip install episia`).

## 6 Research Impact and Field Applications

Episia was developed at Xcept-Health to meet the concrete analytical needs encountered in public health practice in Burkina Faso. Its design takes into account the practical requirements of field epidemiologists.

### 6.1 Outbreak Response: Meningitis in the Sahel

“Lapeyssonnie’s belt” or African meningitis belt, is a region of sub-Saharan Africa particularly vulnerable to meningococcal meningitis epidemics. WHO protocols require weekly monitoring of incidence rates at the district level. Episia’s AlertEngine implements these algorithms, thus enabling the automated identification of “alert” and “epidemic” weeks directly from DHIS2 data. For example, in the event of a suspected outbreak, an analyst can use Episia to extract data from the past 52 weeks, determine the current epidemiological week, and run an SEIR model to forecast the potential peak and duration of the epidemic under different vaccination scenarios.

### 6.2 Vaccine Programme Evaluation

Rigorous biostatistical methods such as the evaluation of large-scale public health interventions are required. Episia offers a suite of validated functions allowing the calculation of a vaccine’s effectiveness from cohort studies or case-control studies, including stratified analyses using the Mantel-Haenszel method to account for confounding factors such as age or geographic region. For example, an analysis can calculate the risk ratio (RR) and its 95

### 6.3 Diagnostic Test Assessment

In many resource-limited settings, the deployment of rapid diagnostic tests (RDTs) for malaria or COVID-19 is critical. Episia’s `diagnostic_test_2×2` and `roc_analysis` functions allow researchers to evaluate the sensitivity, specificity, and predictive values of these tests across different field prevalences. This supports evidence-based procurement decisions at the national or district level.

## 7 Discussion and Future Directions

By offering a high-quality open-source alternative to proprietary software, while lowering the financial barriers to conducting advanced epidemiological analyses, Episia represents a step toward digital sovereignty in the field of public health for African countries.

### 7.1 The “Offline-First” Advantage

One of Episia’s most critical features is its zero network dependency at runtime. Once installed, all modeling, statistical, and reporting functions operate 100

### 7.2 Ease of Use and Reproducibility

Episia’s high-level epi namespace is designed to be approachable for analysts who may not be expert Python developers. By unifying the API, Episia promotes reproducible research : an entire analysis from data ingestion to final report can be captured in a single Jupyter notebook or Python script, making it easy for peers to audit and replicate the results.

### 7.3 Limitations and Future Work

At this stage of development, Episia focuses on deterministic compartmental models. Therefore, to be more efficient, our perspectives include integrating stochastic models to better represent the dynamics of small populations. We also plan to integrate machine learning components for anomaly detection in surveillance data and to extend the DHIS2 client so that it supports “tracking” data (at the individual level) in addition to aggregated data, as well as a suite of spatial modeling.

## 8 Conclusion

Validated and ready for use, Episia, through its comprehensiveness, addresses the challenges of epidemiology in resource-limited settings. By offering a suite that provides modules for advanced modeling and robust biostatistical tools, within a single set of offline-usable features while integrating access to DHIS2 data, this tool enables public health professionals to be more efficient in the face of epidemic threats. Therefore, we invite the global health and digital communities to contribute to this project.

## Ethics Statement

This work involves no collection or use of human subjects data. All analyses are conducted on simulated or publicly available datasets.

## Acknowledgements

Our thanks go to the Xcept-Health team for their support throughout the development as well as the OpenEpi development team for providing a freely accessible reference implementation that allowed the systematic validation of Episia.

