## Supplementary Material for "Episia: An Open-Source Python Library for Epidemiological Surveillance, Modeling, and Biostatistics in Resource-Limited Settings"

#### Overview

This document provides analytical details underlying Figures 3 and 4 of the main manuscript, as requested by the medRxiv screening team. It comprises two sections:

**S1 - Validation Analysis (Figure 4):** complete numerical concordance between Episia and OpenEpi across 12 independent test cases, 25 metrics, and 45 automated checks, with full annotated code.

**S2 - DHIS2 Surveillance Workflow Demonstration (Figure 3):** reproducible end-to-end pipeline using real Episia classes on synthetic meningitis surveillance data from Burkina Faso.

**Source code: https://github.com/Xcept-Health/episia | Zenodo DOI: 10.5281/zenodo.19429374 | PyPI: pip install episia**

### S1. Validation Analysis (Figure 4)

#### S1.1 Methodology

Validation followed a systematic concordance-testing protocol. For each OpenEpi module, reference inputs were taken from OpenEpi published training examples (Sullivan et al., 2009). The same inputs were supplied to the corresponding Episia function. A check passed if the absolute difference |v_Episia - v_OpenEpi| was at most 0.005 or the relative difference was at most 0.5%. All 45 checks passed at Episia v0.1.0.

#### S1.2 Setup

##### Imports and tolerance configuration:

```
from episia.data import SurveillanceDataset, AlertEngine
from episia.stats.contingency import Table2x2, risk_ratio, odds_ratio
from episia.stats.descriptive import proportion_ci, CI_Method, incidence_rate
from episia.stats.diagnostic  import diagnostic_test_2x2
from episia.stats.samplesize  import sample_size_risk_ratio, sample_size_odds_ratio
from episia.stats.stratified  import mantel_haenszel_or

TOL_ABS = 0.005
TOL_PCT = 0.5    # percent

def check(label, episia_val, ref_val, tol=TOL_ABS):
    diff = abs(episia_val - ref_val)
    pct  = diff / max(abs(ref_val), 1e-9) * 100
    ok   = diff <= tol or pct <= TOL_PCT
    print(f"  [{'PASS' if ok else 'FAIL'}]  {label:40s}  "
          f"episia={episia_val:.5f}  ref={ref_val:.5f}  delta={diff:.5f}")
    return ok
```

#### S1.3 Test Cases

##### Test 1: Two-by-Two cohort table (Emory/OpenEpi training)

```
# Cases: exposed=84, unexposed=29 | Non-cases: exposed=39, unexposed=64
t1   = Table2x2(a=84, b=29, c=39, d=64)
rr1  = t1.risk_ratio()
or1  = t1.odds_ratio()
chi1 = t1.chi_square(correction=False)

# Output: RR=2.1901 (1.5823-3.0313) | OR=4.7533 (2.6606-8.4919) | chi2=29.2352 p<0.0001
check("RR",        rr1.estimate,   2.19010)  #   PASS   delta=0.00002
check("RR CI lower", rr1.ci_lower,  1.58230)  # PASS  delta=0.00001
check("RR CI upper", rr1.ci_upper,  3.03130)  # PASS  delta=0.00001
check("OR",        or1.estimate,   4.75330)  #   PASS   delta=0.00002
check("OR CI lower", or1.ci_lower,  2.66060)  # PASS  delta=0.00000
check("OR CI upper", or1.ci_upper,  8.49190)  # PASS  delta=0.00000
```

##### Test 2: SSCohort tutorial (OR=2.667)

```
# OpenEpi SSCohort tutorial: p_exposed=40%, p_unexposed=20%
t2  = Table2x2(a=40, b=20, c=60, d=80)
rr2 = t2.risk_ratio()
or2 = t2.odds_ratio()

check("Risk exposed",   t2.risk_exposed,    0.4000)  # PASS  delta=0.00000
check("Risk unexposed", t2.risk_unexposed,  0.2000)  # PASS  delta=0.00000
```

```
check("RR",              rr2.estimate,       2.0000)  # PASS  delta=0.00000
check("OR",              or2.estimate,       2.6667)  # PASS  delta=0.00003
```

##### Test 3: Risk difference and attributable fraction

```
t3   = Table2x2(a=84, b=29, c=39, d=64)
rd3  = t3.risk_difference()
af_e = t3.attributable_fraction_exposed()
paf  = t3.attributable_fraction_population()

# Output: RD=0.3711 (0.2461-0.4961) | AF_exposed=54.3% | PAF=46.9%
check("Risk difference", rd3['estimate'], 0.37110)  # PASS  delta=0.00000
check("RD  CI  lower",   rd3['ci_lower'],  0.24610)  #  PASS  delta=0.00001
check("RD  CI  upper",   rd3['ci_upper'],  0.49610)  #  PASS  delta=0.00001
check("AF exposed",     af_e,              0.54340)  #  PASS  delta=0.00001
check("PAF population",  paf,              0.46940)  #  PASS  delta=0.00000
```

##### Test 4: Fisher exact test (small sample)

```
# Small table where chi-square is inappropriate
t4  = Table2x2(a=5, b=0, c=1, d=4)
fe4 = t4.fisher_exact()

# Output: Fisher p=0.047619 (OpenEpi ref: 0.0476)
check("Fisher exact p", fe4['p_value'], 0.047619)  # PASS  delta=0.00000
```

##### Test 5: Proportion Wilson CI (k=7, n=98)

```
p5 = proportion_ci(k=7, n=98, method=CI_Method.WILSON)

# Output: p=0.071429 | CI (0.035028 - 0.140160)
check("Proportion",   p5.proportion,  0.071429)  #  PASS  delta=0.00000
check("Wilson  CI  lower", p5.ci_lower,  0.03497)  #  PASS  delta=0.00006
check("Wilson CI upper", p5.ci_upper,   0.14015)   # PASS  delta=0.00001
```

##### Test 6: Proportion - five CI methods (k=45, n=200)

```
# OpenEpi Proportion module - all five methods
methods = [CI_Method.WILSON, CI_Method.WALD, CI_Method.CLOPPER_PEARSON,
           CI_Method.JEFFREYS, CI_Method.AGRESTI_COULL]

# All five methods return p=0.225 exactly
for m in methods:
    r = proportion_ci(k=45, n=200, method=m)
    check(f"p == 0.225 ({m.value})", r.proportion, 0.2250)  # PASS x5

# Wilson:          CI (0.17262 - 0.28774)
# Clopper-Pearson: CI (0.16910 - 0.28924)
# Jeffreys:        CI (0.17134 - 0.28655)
```

##### Test 7: Incidence rate with Byar CI (cases=4, person-years=1000)

```
ir = incidence_rate(cases=4, person_time=1000)

# Output: rate=0.004000 | Byar CI (0.001090 - 0.010242)
check("Incidence  rate",  ir['rate'],    0.004000)  #  PASS  delta=0.00000
check("Byar CI lower",  ir['ci_lower'], 0.001090)  # PASS  delta=0.00000
check("Byar CI upper",  ir['ci_upper'], 0.010242)  # PASS  delta=0.00000
```

##### Test 8: Sample size cohort study (OpenEpi SSCohort)

```
# Parameters: p0=20%, RR=2.0, alpha=0.05, power=80%
ss8 = sample_size_risk_ratio(rr_expected=2.0, p0=0.20, alpha=0.05, power=0.80)

# Output: 82 per group, total 164
```

```
check("n per group", ss8.n_per_group, 82)   # PASS  delta=0.00000
check("n total",     ss8.n_total,    164)   # PASS  delta=0.00000
```

##### Test 9: Sample size case-control (OpenEpi SSCaseControl)

```
# Parameters: OR=2.5, p0=0.30, alpha=0.05, power=80%
ss9 = sample_size_odds_ratio(
    proportion_exposed_controls=0.30,
    odds_ratio=2.5, power=0.80, alpha=0.05)

# Output: 80 cases, 80 controls, total 160
check("n cases",    ss9.n_cases,    80)   # PASS  delta=0.00000
check("n controls", ss9.n_controls, 80)   # PASS  delta=0.00000
check("n total",    ss9.n_total,   160)   # PASS  delta=0.00000
```

##### Test 10: Diagnostic test evaluation (OpenEpi Screening)

```
# TP=80, FP=10, FN=20, TN=90 (200 patients total)
diag = diagnostic_test_2x2(tp=80, fp=10, fn=20, tn=90)

check("Sensitivity",  diag.sensitivity, 0.8000)   # PASS  delta=0.00000
check("Specificity",  diag.specificity, 0.9000)   # PASS  delta=0.00000
check("PPV",          diag.ppv,         0.88889)  # PASS  delta=0.00000
check("NPV",          diag.npv,         0.81818)  # PASS  delta=0.00000
check("LR+",          diag.lr_positive, 8.0000)   # PASS  delta=0.00000
check("Accuracy",     diag.accuracy,    0.8500)   # PASS  delta=0.00000
check("Youden index", diag.youden,      0.7000)   # PASS  delta=0.00000
```

##### Test 11: Mantel-Haenszel stratified OR

```
# Source: OpenEpi 2x2xK stratified analysis module
strata = [
    Table2x2(a=18, b=8,  c=26, d=22),   # age group 1
    Table2x2(a=12, b=4,  c=5,  d=10),   # age group 2
]
mh = mantel_haenszel_or(strata)

check("MH pooled OR", mh.common_or,  2.66900)  # PASS  delta=0.00048
check("MH CI lower", mh.or_ci[0],  1.38600)  # PASS  delta=0.00018
check("MH CI upper", mh.or_ci[1],   5.13700)  # PASS  delta=0.00013
```

##### Test 12: Population Attributable Fraction (PAF)

```
t12  = Table2x2(a=84, b=29, c=39, d=64)
paf12 = t12.attributable_fraction_population()
pe12  = t12.a / t12.total_cases    # proportion exposed among cases = 0.7434

# PAF = Pe*(RR-1) / (Pe*(RR-1)+1) = 0.4694
check("PAF", paf12, 0.46940)  # PASS  delta=0.00000
```

#### S1.4 Complete Concordance Results (Table S1)

All 25 metrics across 12 test cases. All 45 automated checks passed at tolerance |delta| <= 0.005 or relative <= 0.5%. Episia v0.1.0.

| Module | Metric | OpenEpi ref | Episia | \|delta\| | Status |
|---|---|---|---|---|---|
| 2x2 Table | RR (point est.) | 2.19010 | 2.19008 | 0.00002 | PASS |
| | RR CI lower | 1.58230 | 1.58231 | 0.00001 | PASS |
| | RR CI upper | 3.03130 | 3.03129 | 0.00001 | PASS |

| Module | Metric | OpenEpi ref | Episia | \|delta\| | Status |
|---|---|---|---|---|---|
| | OR (point est.) | 4.75330 | 4.75332 | 0.00002 | PASS |
| | OR CI lower | 2.66060 | 2.66060 | 0.00000 | PASS |
| | OR CI upper | 8.49190 | 8.49190 | 0.00000 | PASS |
| | Risk difference | 0.37110 | 0.37110 | 0.00000 | PASS |
| | AF (exposed) | 0.54340 | 0.54339 | 0.00001 | PASS |
| | PAF (population) | 0.46940 | 0.46940 | 0.00000 | PASS |
| Fisher | p-value | 0.04762 | 0.04762 | 0.00000 | PASS |
| Proportions | Wilson CI lower | 0.03497 | 0.03503 | 0.00006 | PASS |
| | Wilson CI upper | 0.14015 | 0.14016 | 0.00001 | PASS |
| Incidence | Rate (Byar) | 0.00400 | 0.00400 | 0.00000 | PASS |
| | Byar CI lower | 0.00109 | 0.00109 | 0.00000 | PASS |
| | Byar CI upper | 0.01024 | 0.01024 | 0.00000 | PASS |
| Sample size | n per group | 82.000 | 82.000 | 0.00000 | PASS |
| | n total | 164.000 | 164.000 | 0.00000 | PASS |
| Diagnostic | Sensitivity | 0.80000 | 0.80000 | 0.00000 | PASS |
| | Specificity | 0.90000 | 0.90000 | 0.00000 | PASS |
| | PPV | 0.88889 | 0.88889 | 0.00000 | PASS |
| | NPV | 0.81818 | 0.81818 | 0.00000 | PASS |
| | LR+ | 8.00000 | 8.00000 | 0.00000 | PASS |
| | Accuracy | 0.85000 | 0.85000 | 0.00000 | PASS |
| | Youden index | 0.70000 | 0.70000 | 0.00000 | PASS |
| Mantel-Haenszel | Pooled OR | 2.66900 | 2.66852 | 0.00048 | PASS |
| | MH CI lower | 1.38600 | 1.38618 | 0.00018 | PASS |
| | MH CI upper | 5.13700 | 5.13713 | 0.00013 | PASS |

*Table S1. Numerical concordance between Episia v0.1.0 and OpenEpi. OpenEpi reference values from Sullivan et al. (2009) training examples. Validation notebook: github.com/Xcept-Health/episia/blob/main/exemples/episia_vs_openepi.ipynb*

### S2. DHIS2 Surveillance Workflow Demonstration (Figure 3)

#### S2.1 Context

Figure 3 is an architectural diagram of the Episia data pipeline. This section instantiates every component of that diagram using meningitis_2024.csv, a synthetic weekly meningitis surveillance dataset for the Centre district of Burkina Faso (population 500,000, 52 weeks). All code uses the public Episia API as documented at github.com/Xcept-Health/episia. Executed at Episia v0.1.0.

#### S2.2 Full Demonstration Code and Output

##### Step 1: Data ingestion (SurveillanceDataset):

```
from episia.data import SurveillanceDataset, AlertEngine
from episia.viz.curves import plot_epicurve

# In production: replace from_csv() with DHIS2Client.get_data_values()
dataset = SurveillanceDataset.from_csv(
    "meningitis_2024.csv",
    date_col="date",      cases_col="cases",
    deaths_col="death",   district_col="district",
    population_col="population",
)
print(dataset.summary())
```

##### Output:

```
{
  'n_records': 52,  'total_cases': 539,
  'date_start': '2024-01-07',  'date_end': '2024-12-29',
  'total_deaths': 51,  'cfr': 0.09462,
  'n_districts': 1,  'districts': ['Centre']
}
```

##### Step 2: Alert detection (AlertEngine.run):

```
engine  = AlertEngine(dataset)
alerts  = engine.run(threshold=10, zscore_threshold=2.0)
summary = engine.alert_summary(alerts)
print(summary)
```

##### Output:

```
{
  'n_alerts': 23,
  'severity_counts': {'alert': 11, 'epidemic': 9, 'warning': 3},
  'first_alert': '2024-02-05T00:00:00.000000000',
  'last_alert':  '2024-10-14T00:00:00.000000000'
}
```

##### Step 3: Individual alert output:

```
for a in alerts:
    print(a.period, a.severity, a.message)

# 2024-02-05  alert    12 cas >= seuil 10
# 2024-02-12  alert    18 cas >= seuil 10
# 2024-02-19  epidemic 22 cas >= seuil 10
# 2024-02-26  epidemic 28 cas >= seuil 10
# 2024-03-04  epidemic 35 cas >= seuil 10
# 2024-03-04  warning  Z-score=2.48 >= 2.0
# 2024-03-11  epidemic 42 cas >= seuil 10
```

```
# 2024-03-11  epidemic Z-score=3.18 >= 2.0
# 2024-03-18  epidemic 38 cas >= seuil 10
# 2024-03-25  epidemic 31 cas >= seuil 10
# 2024-04-01  epidemic 25 cas >= seuil 10
# 2024-09-23  epidemic 20 cas >= seuil 10
# ...  (23 alerts total, see Table S1 for full list)
```

**Step 4: Visualization via episia.viz.curves.plot_epicurve():**

```
agg    = dataset.aggregate(freq="W")
times  = agg["period"].dt.strftime("%Y-W%W").values
values = agg[dataset.cases_col].values.astype(float)

fig = plot_epicurve(
    times=times, values=values,
    title="Meningitis Surveillance - Centre District, Burkina Faso 2024",
    xlabel="Epidemiological week",
    ylabel="Cases",
    backend="matplotlib",
    theme="scientific",
)
fig.savefig("figure3_dhis2_surveillance.png", dpi=300, bbox_inches="tight")
```

#### S2.3 Interpretation

The AlertEngine correctly identified two epidemic waves in 2024: a primary wave in February-April (peak: 42 cases, week 11) and a secondary wave in September-October (peak: 20 cases, week 39), consistent with the bimodal meningococcal pattern in the Sahel. Of the 23 alert-weeks, 9 crossed the epidemic threshold (>= 2x the alert threshold) and 3 were flagged independently by Z-score (>= 2.0). CFR was 9.5%, within the range reported for meningococcal meningitis in Burkina Faso. This demonstrates that the workflow in Figure 3 runs end-to-end with real Episia code.

---

World Health Organization. Epidemic meningitis surveillance in the African meningitis belt. Geneva: WHO; 2014. Report No.: WHO/HSE/PED/CED/14.1.
